# Clinicogenomic landscape and function of *PIK3CA*, *AKT1*, and *PTEN* mutations in breast cancer

**DOI:** 10.1101/2025.06.18.25329632

**Authors:** Jacqueline J. Tao, Saumya D. Sisoudiya, Hanna Tukachinsky, Alexa Schrock, Smruthy Sivakumar, Ethan S. Sokol, Neil Vasan

## Abstract

**Purpose:** To comprehensively characterize the clinical and genomic landscapes of *PIK3CA, AKT1,* and *PTEN* alterations and examine their functional implications in AKT-driven breast cancer.

**Experimental Design:** Comprehensive genomic profiling of 51,767 breast tumors was performed with FoundationOne^®^CDx or FoundationOne^®^. We examined the genomic landscape of *PIK3CA*, *PTEN*, and *AKT1* alterations and their distribution across clinical variables of interest. Prior deep mutational scanning (DMS) data was used to functionally characterize clinical *PTEN* variants.

**Results:** There were 29,157 total variants across *PIK3CA, AKT1,* and *PTEN*, including pathogenic variants and VUS. The most frequently altered gene was *PIK3CA* (37.4% of cases), followed by *PTEN* (13.5%), then *AKT1* (5.4%). The most common alterations in each gene were *PIK3CA* H1047R (35.6% of *PIK3CA*-altered cases), E545K (19.7%), and E542K (11.7%); *AKT1* E17K (69.7%); and *PTEN* homozygous copy number deletion (37.3%). *PIK3CA* alterations were less prevalent in patients of African genetic ancestry (27.1% vs 38.6% in European genetic ancestry), while *AKT1* and *PTEN* alterations were balanced across ancestries. *PIK3CA*, *AKT1*, and *PTEN* pathogenic alterations were all mutually exclusive to each other. Using available DMS data on missense *PTEN* mutations, we found that 32.5% showed discordant effects on protein stability and phosphatase activity, underscoring the need for functional validation beyond predicted loss-of-function.

**Conclusions:** Here we present the landscape of *PIK3CA*, *AKT1*, and *PTEN* alterations in the largest clinical cohort examined to date. The functional implications of lesser-known variants in each gene warrant further investigation by tools such as deep mutational scanning.

## Introduction

The AKT pathway plays a critical role in multiple cellular functions including growth, proliferation, survival, and metabolism, and is one of the most frequently altered pathways across cancers. *PIK3CA* encodes the catalytic subunit of PI3Kα, which recruits and activates AKT and subsequent downstream signaling through mTORC1^1^. The lipid phosphatase PTEN metabolizes the lipid phosphoinositide species PIP_3_, antagonizing the effects of PI3K. Consequently, aberrant PI3K pathway activation primarily arises due to activating mutations in *PIK3CA,* along with activating mutations in *AKT1* and inactivating alterations in *PTEN*^2^.

The AKT pathway is altered in over 40% of estrogen receptor-positive (ER+) breast cancers^3^, and numerous agents targeting pathway constituents have been developed and approved over the last two decades. One of the first approved agents targeting this pathway was the mTOR inhibitor everolimus, which was shown in the BOLERO-2 trial to improve progression free survival (PFS) in combination with exemestane in patients with endocrine-resistant metastatic breast cancer (MBC)^4^. Subsequently, the PI3Kα-specific inhibitor alpelisib was approved for *PIK3CA*-mutated MBC based on the SOLAR-1 trial, where it doubled PFS in combination with fulvestrant^5^. More recently, the mutant-selective PI3Kα inhibitor inavolisib was developed and found to have synergistic activity with palbociclib plus fulvestrant for patients with advanced breast cancer relapsed within 12 months after completing adjuvant endocrine therapy^6^. However, several of these agents are associated with significant on-target toxicities including grade 3+ hyperglycemia in more than one-third of patients receiving alpelisib. Recently, the AKT inhibitor capivasertib was approved for patients with ER+ MBC with mutations in *PIK3CA*, *AKT1*, and/or *PTEN* after progression on first-line endocrine-based therapy^7^. In the CAPItello-291 study, capivasertib combined with fulvestrant more than doubled PFS in patients with qualifying genomic alterations (median PFS 7.3 months versus 3.1 months with fulvestrant alone)^8^. Notably, clinical endpoints were not significantly improved in the wild-type population, highlighting the importance of *PIK3CA*/*AKT1*/*PTEN* alterations as biomarkers of sensitivity to capivasertib. Capivasertib also demonstrated a favorable toxicity profile compared with older generations of PI3K inhibitors, including lower rates of hyperglycemia (2.3% grade 3-4 versus 36.6% grade 3-4 with alpelisib), accelerating its adoption in the clinical setting over older agents.

Although significant progress has been made in characterizing the AKT pathway at the genomic level^9,10^, many prior dedicated studies of *PIK3CA/AKT1/PTEN* have had limited sequencing coverage, analyzed smaller genomic data sets, or examined restricted clinical subsets. An early landscape analysis of *PIK3CA/AKT1/PTEN* used mass spectroscopy-based sequencing to analyze 547 breast tumors^11^; more recent examples have described this 3-gene landscape in 1,967 luminal type breast cancers that underwent gene panel sequencing^12^ and 7,450 hormone receptor-positive breast cancers that underwent next generation sequencing^13^.

With the recent FDA approval of capivasertib for patients with breast cancer harboring *PIK3CA*, *AKT1*, or *PTEN* alterations, it is increasingly critical to understand the clinical landscape of these genomic subsets. We have previously leveraged large-scale genomic data sets to investigate the landscape of multi-*PIK3CA* mutated tumors, enabling unique insights into the genetic heterogeneity and tissue specificities of multi-*PIK3CA* mutations^14^. While *PIK3CA* mutations have been extensively studied, far less is known about the prevalence, co-alteration patterns, and clinical outcomes associated with *AKT1* and *PTEN* alterations in breast cancer. Here we assess the frequencies, distributions, clinical associations, and co-occurrence patterns of *PIK3CA, AKT1,* and *PTEN* variants in the largest breast cancer dataset to date (n = 51,767) that integrates genomic and clinical data across all three alterations, offering a unique opportunity to define their distinct and overlapping roles in therapeutic response and resistance.

## Methods

### Comprehensive genomic profiling of a breast cancer cohort

Comprehensive genomic profiling (CGP) of formalin-fixed, paraffin-embedded tissue sections was performed using the FoundationOne®CDx assay or the laboratory-developed test predecessor, FoundationOne®, as part of routine clinical care in a Clinical Laboratory Improvement Amendments-certified, College of American Pathologists-accredited laboratory (Foundation Medicine, Inc.). This study included 51,767 breast cancer samples profiled between August 2014 and September 2023. Approval for this study, including a waiver of informed consent and Health Insurance Portability and Accountability Act (HIPAA) waiver of authorization, was obtained from the Western Institutional Review Board (protocol no. 20152817).

### Genomic analysis

CGP was performed on hybrid capture-selected libraries spanning exons of at least 300 cancer-related genes and select introns of genes frequently rearranged in cancer. Processing of the sequence data and identification of different classes of genomic alterations was carried out as previously described^15,16^. The alterations identified included single nucleotide variants (missense, nonsense, and splice site mutations), indels (in-frame and frameshift mutations), copy number alterations, and rearrangement events. Alterations were classified as either known or likely pathogenic using annotations from various sources, including their occurrence in the COSMIC database, previous scientific literature-based knowledge about the affected gene (such as truncations and deletions in known tumor suppressor genes), or mutations that have previously been characterized as pathogenic; all other uncharacterized alterations were classified as variants of unknown significance (VUS).

### Histopathologic and clinical characteristics

HER2 status was determined from the HER2 (*ERBB2*) amplification status based on the FoundationOne® or FoundationOne®CDx assays. ER status was derived from accompanying pathology reports, where available. Clinical data including biopsy site (local versus lymph node vs metastatic) were obtained from the submitted test requisition form. Genetic ancestry of each patient was predicted using a single nucleotide polymorphism (SNP)-based approach. Briefly, the profiled SNPs that overlapped with those captured in phase 3 of the 1000 Genomes Project were projected down to five principal components that were then used to train a random forest classifier to identify the following ancestry groups: European (EUR), African (AFR), East Asian (EAS), South Asian (SAS), and admixed American (AMR)^17^.

### Co-occurrence and mutual exclusivity analyses

The dataset was interrogated for patterns of co-occurrence and mutual exclusivity of *PIK3CA, AKT1,* and *PTEN* mutations with other cancer-associated genes targeted as part of the assays. A Fisher’s exact test was used to identify significantly co-occurring (OR > 1) as well as mutually exclusive (OR < 1) patterns using a FDR-adjusted p-value threshold of 0.05. A Benjamini-Hochberg FDR correction was applied to adjust for multiple comparisons.

### Functional characterization of clinical PTEN alterations

To explore the functional implications of the *PTEN* variants in this clinical cohort, we examined two published deep mutational scanning (DMS) data sets characterizing the effects of single amino acid substitutions on *PTEN* function. In Matreyek et al 2018^18^, abundance scores were determined by measuring the steady-state intracellular abundance of *PTEN* variants in cultured human cells using VAMP-seq (Variant Abundance by Massively Parallel Sequencing). Abundance scores were refined and generated for additional *PTEN* variants in Matreyek et al 2021^19^. Scores ranged from 0, reflecting total loss of abundance, to 1, reflecting wild type (WT)-like abundance. Based on the abundance score, each *PTEN* variant was assigned an abundance class of low, possibly low, possibly WT-like, or WT-like. In Mighell et al 2018^20^, fitness scores reflecting the lipid phosphatase activity of *PTEN* variants were determined by assessing enzymatic ability to reverse toxicity via PIP_3_ dephosphorylation in a humanized yeast system. Variants were assigned functional categories based on fitness score percentiles: scores below the upper two-tailed 95^th^ percentile for nonsense variants (≤ -2.13) were designated truncation-like, scores within the two-tailed 95^th^ percentile for synonymous variants (-1.11 to 0.89) were designated WT-like, and scores in between these ranges (-2.13 to -1.11) were designated hypomorphic. Here we extracted abundance and fitness scores corresponding to the *PTEN* missense and nonsense variants in our clinical cohort and assigned functional classifications for each variant based on these scores. Functional designations created in the original publications were consolidated into binary categories of pathogenic (abundance classes of low and possibly low; fitness score categories of truncation-like and hypomorphic) and non-pathogenic (abundance classes of WT-like and possibly WT-like; fitness score categories of WT-like and >WT). For each missense *PTEN* variant, functional classifications derived from the abundance score and fitness score were compared.

### Statistical analyses

Categorical variables were compared between groups using the Fisher’s exact test, with an FDR correction for multiple testing. A p-value ≤0.05 was considered statistically significant. Statistics, computation, and plotting were carried out using R 3.6.1 (R Foundation for Statistical Computing).

### Data availability

All relevant data are provided within the article and its accompanying Supplementary Data. Due to HIPAA requirements, we are not authorized to share individualized patient genomic data, which contains potentially identifying or sensitive patient information. Foundation Medicine is committed to collaborative data analysis, with well-established and widely utilized mechanisms by which investigators can query our core genomic database of >700,000 de-identified sequenced cancers to obtain aggregated datasets. For more information and mechanisms of access, please contact the corresponding author(s) or the Foundation Medicine, Inc. Data Governance Council at.

## Results

### Baseline cohort characteristics

This analysis included 51,767 breast cancer tumor samples. The majority (51,199; 99%) of the cohort were female and median age was 59 years (IQR 50-68 years). HER2 amplification was assessed in all samples, of which 4312 (8.3%) were HER2+. ER status was available in 4219 cases, of which 2409 (57.1% of assessable cases) were ER+/HER2- and 1408 (33.4%) were ER-/HER2-. The most common ancestries represented were European (72%) and African (14%). Tumor samples were obtained from a metastatic site (47%), local site (33%), or lymph node (12%), with the remaining samples having unknown or ambiguous biopsy site information (8%).

### Prevalence and spectrum of AKT pathway alterations

There were 29,157 total variants in *PIK3CA, AKT1,* and *PTEN* identified in the cohort. The most frequently altered PI3K-pathway gene was *PIK3CA* (37.4% of samples; 19,384/51,767), followed by *PTEN* (13.5%; 6,991/51,767) then *AKT1* (5.4%; 2,782/51,767), including both pathogenic alterations and VUS in all genes. When taking only pathogenic alterations into account, *PIK3CA* was altered in 36.5% of all samples, *PTEN* in 12.7%, and *AKT1* in 4.7%. Among 1,024 distinct *PIK3CA* alterations identified, the most common variant type was missense mutations (690/1,024; 67.4% of all *PIK3CA* variants) followed by in-frame mutations (252; 24.6%; Figure 1, Supplementary Table S1). The most common mutations were H1047R (35.6% of *PIK3CA*-altered cases), E545K (19.7%), and E542K (11.7%), consistent with the known mutational hotspots^21^. The fourth most common *PIK3CA* alteration was amplification (6.1%). Among 288 distinct *AKT1* alterations identified, missense mutations remained the most common variant type (205/288; 71.2% of all *AKT1* variants), with the most common mutation being E17K (69.7% of *AKT1*-altered cases; Figure 1, Supplementary Table S2). The second most common *AKT1* alteration was gene amplification (14.0%). Finally, among 1,723 distinct *PTEN* alterations identified, the most common variant types were frameshift mutations (721/1,723; 41.9% of *PTEN* variants), missense mutations (494; 28.7%), and splice site mutations (246; 14.3%). The most common individual *PTEN* alteration was gene deletion (37.3% of *PTEN*-altered cases; Figure 1, Supplementary Table S3).

**Figure 1.**
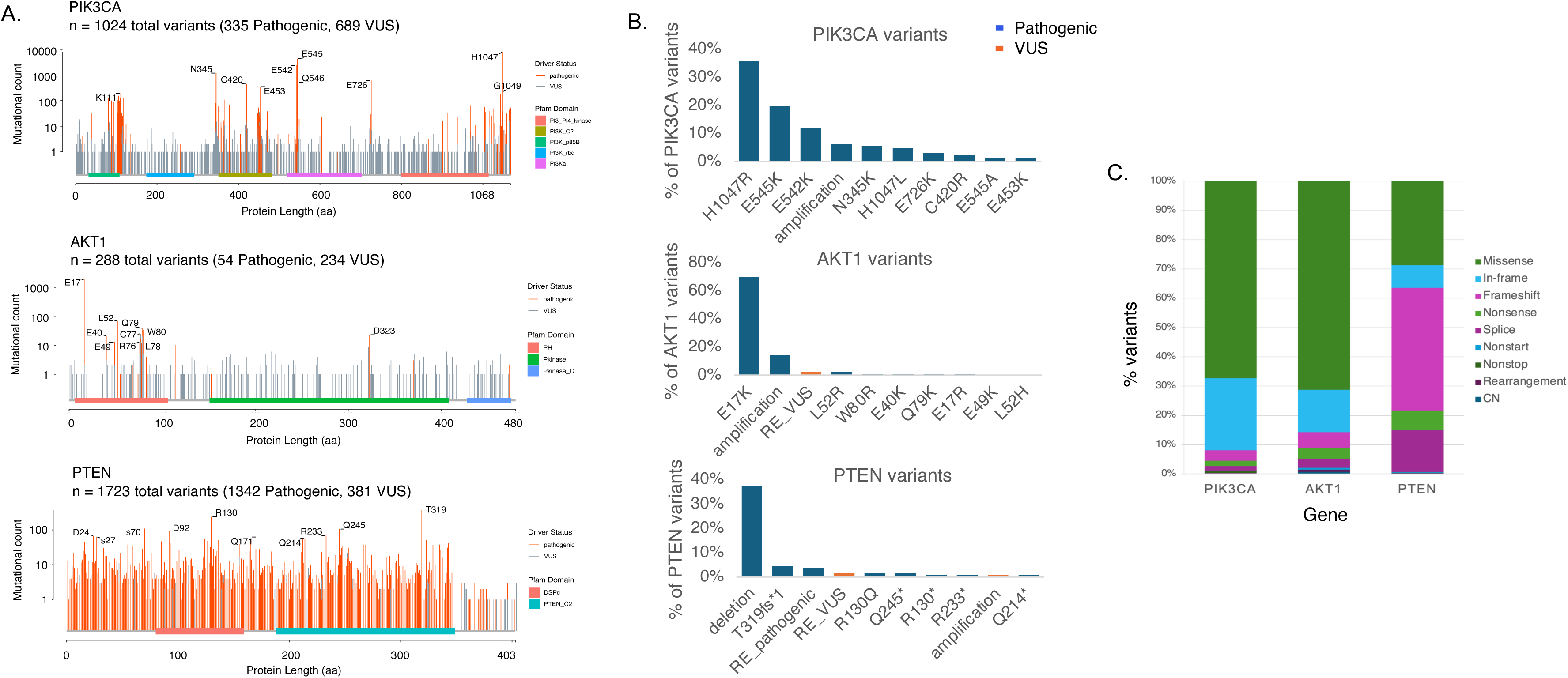
Frequency, distribution, and coding type of *PIK3CA*, *AKT1*, and *PTEN* variants. The most frequently altered PI3K-pathway gene was *PIK3CA* (37.4% of patients), followed by *PTEN* (13.5%) then *AKT1* (5.4%). **A)** Lollipop plots of *PIK3CA*, *AKT1*, and *PTEN* mutations. **B)** Prevalence of individual variants in each gene. The most common variants in each gene were *PIK3CA* H1047R (35.6% of *PIK3CA*-altered cases), *AKT1* E17K (69.7%), and *PTEN* deletion (37.3%). **C)** Distribution of variant coding types in each gene. The most common variant coding types were *PIK3CA* missense mutation (67.4%), *AKT1* missense mutation (71.2%), and *PTEN* frameshift mutation (41.9%).

Multiple pathogenic *PIK3CA* alterations were seen in 3,148 samples (16.2% of all *PIK3CA*-altered samples and 6.1% of overall cohort). Pathogenic alterations in more than one gene were most commonly seen with *PIK3CA* and *PTEN* (2018 samples, 4% of cohort) followed by *PIK3CA* and *AKT1* (322 samples, 0.6%) then *AKT1* and *PTEN* (71 samples, 0.14%).

### Clinical and demographic associations

We next examined the distribution of *PIK3CA, AKT1,* and *PTEN* mutations stratified by clinical variables including receptor subtype (Figures 2A and 2D, Supplementary Table S4), patient ancestry (Figures 2B and 2E, Supplementary Table S5), and biopsy site (Figures 2C and 2F, Supplementary Table S6). *PIK3CA* pathogenic variants were more prevalent in ER+/HER2- and HER2+ disease (40.3%, *p*=6.0x10^-36^ and 37.6% of cases, *p*=2.3x10^-32^) than in ER-/HER2- disease (20.9%), consistent with prior reports^11,22^. However, variants outside the 542, 545, and 1047 codons were more common in ER-/HER2- (42%) than ER+/HER2- or HER2+ cases (33%, *p*=0.002 and 33%, *p*=0.002). *AKT1* mutations were most prevalent in ER+/HER2- disease (6.0%) and less common in HER2+ or ER-/HER2- disease (1.7%, *p*=4.7x10^-20^ and 3.0%, *p*=3.5x10^-5^). In contrast, *PTEN* alterations were more commonly found in ER-/HER2- (17.9%) than in ER+/HER2- and HER2+ disease (11.3%, *p*=1.8x10^-8^ and 3.9%, *p*=2x10^-57^).

**Figure 2.**
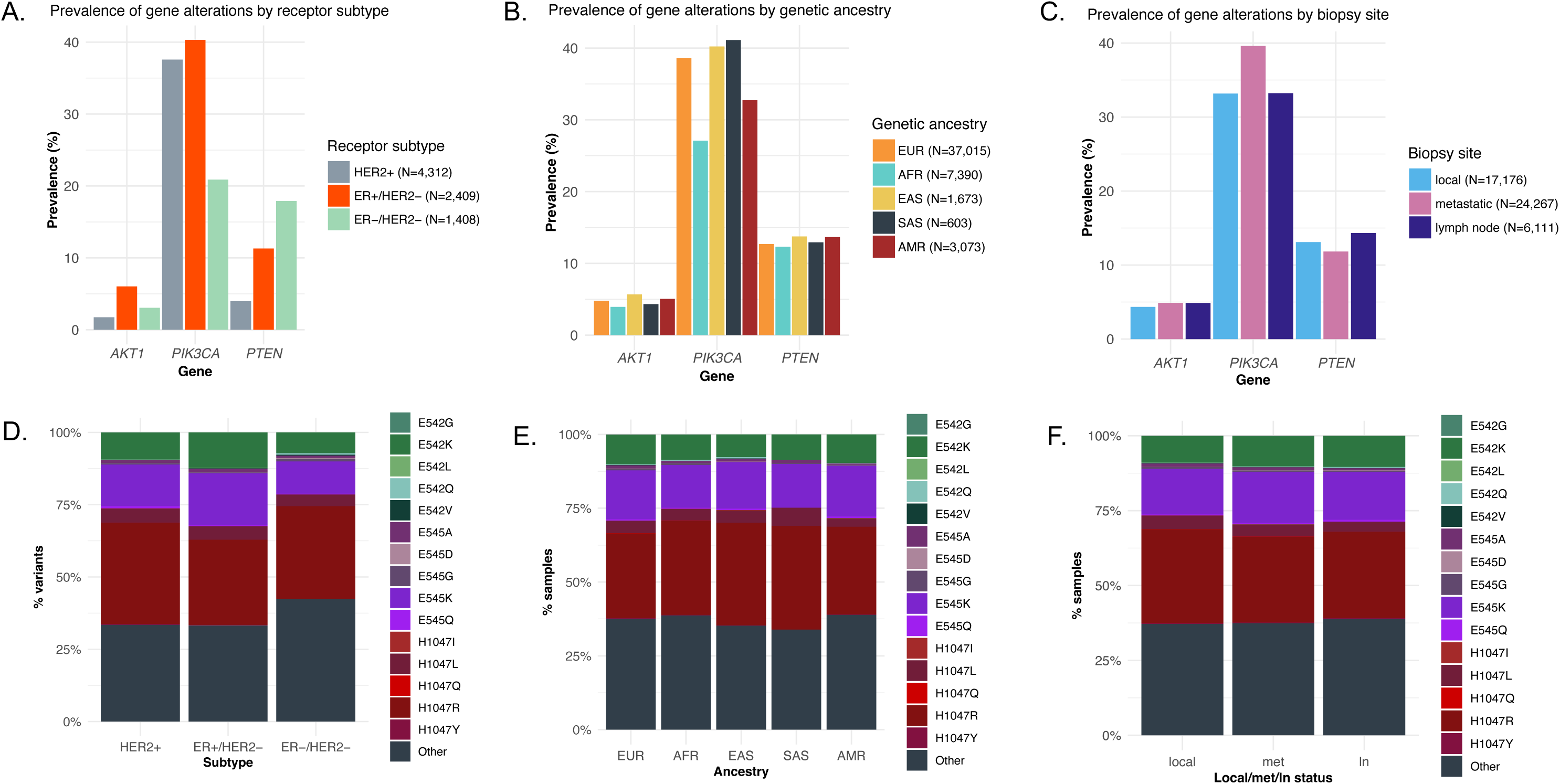
Prevalence of gene variants by breast cancer receptor subtype, genetic ancestry, and biopsy site. **A)** *PIK3CA* variants were enriched in ER+/HER2- and HER2+ disease (altered in 40.3% and 37.6% of cases, respectively) and *AKT1* variants were enriched in ER+/HER2-disease (6.0%). In contrast, *PTEN* variants were most prevalent in ER-/HER2- disease (17.9%). **B-C)** *PIK3CA* alterations were less prevalent in patients with African ancestry (27.1% vs 38.6% in European ancestry) and enriched in metastases (39.6% vs 33.2% in local tumors). **D)** *PIK3CA* variants outside the 542, 545, and 1047 codons were more common in ER-/HER2- disease (42%) compared to ER+/HER2- or HER2+ disease (33% and 33%). **E, F)** *PIK3CA* variants by ancestry and biopsy site.

*PIK3CA* pathogenic alterations were less prevalent in patients with African ancestry than other groups, both in the full cohort (27.1% vs 38.6% in Europeans, *p*=9.6x10^-81^) and in the ER+/HER2- disease subset (31.2% vs 41.1% in Europeans, *p*=0.002). Additionally, among ER+/HER2- cases only, patients with South Asian ancestry had a higher prevalence of *PIK3CA* pathogenic alterations than other groups (68.9% vs 41.1% in Europeans, *p*=0.002). *PIK3CA* pathogenic variants were enriched in metastases compared to local tumors (39.6% vs 33.2%, *p*=6x10^-41^). The distribution of individual *PIK3CA* variants was similar across ancestries and biopsy sites. The prevalence of pathogenic *AKT1* and *PTEN* alterations was similar across all ancestries and biopsy sites.

### Co-occurrence and mutual exclusivity analyses

Next, we characterized the patterns of co-occurrence and mutual exclusivity between pathogenic variants in *PIK3CA, AKT1,* and *PTEN* and other sequenced genes (Figure 3; Supplementary Tables S7-S9). Regarding PI3K pathway genes, *PIK3CA*, *AKT1*, and *PTEN* pathogenic alterations were all strongly mutually exclusive with each other (*PIK3CA/AKT1* odds ratio [OR] 0.25, *p*<10^-60^; *PIK3CA/PTEN* OR 0.74, *p*=6.6x10^-25^; *AKT1/PTEN* OR 0.20, *p*<10^-60^), suggesting that a single driver mutation may be sufficient for aberrant pathway activation. Notably, the degree of mutual exclusivity was lowest between *PIK3CA* and *PTEN*, corresponding to the higher rates of co-mutations observed in these two genes compared to the other combinations.

**Figure 3.**
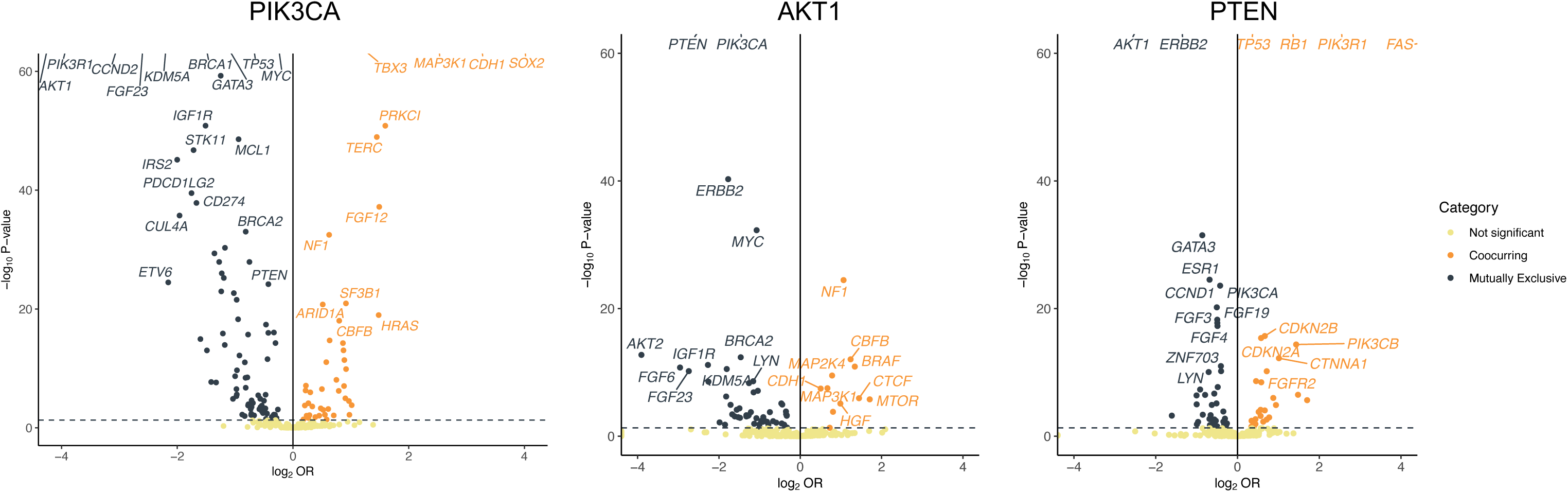
Co-occurrence and mutual exclusivity analyses of pathogenic variants in *PIK3CA*, *AKT1*, and *PTEN*. Pathogenic variants in these 3 genes were all mutually exclusive to each other. Notable co-occurrences included *PIK3CA* with *SOX2, PRKCI, MAP3K1, TERC, CDH1,* and *TBX3* ; *AKT1* with *NF1* and *CDH1*; and *PTEN* with *FAS*, *TP53*, and *RB1*. Notable mutual exclusivities include *PIK3CA* with *TP53, BRCA1/2,* and *STK11* ; *AKT1* with *ERBB2* and *MYC;* and *PTEN* with *ERBB2, GATA3,* and *ESR1.* Y-axis is capped at p-value of 10E-60.

*PIK3CA* was significantly co-altered with several other cancer-associated genes including *SOX2, PRKCI, MAP3K1, TERC, CDH1,* and *TBX3*. Interestingly the strongest association was with *SOX2*, a relationship that has not previously been described in the breast cancer literature, though there are reports suggesting a role of *PIK3CA* and *SOX2* co-amplification in the pathogenesis of head and neck cancers^23^. Several other significantly co-occurring genes have well-known associations with *PIK3CA* including *CDH1* and *TBX3*, whose losses are hallmarks of invasive lobular carcinoma (ILC)^24,25^, and *MAP3K1*, which is associated with luminal A phenotype^26–28^. These findings are consistent with the enrichment of ILC and luminal A breast cancers for *PIK3CA* mutations. Several tumor suppressors were mutually exclusive with *PIK3CA* including *TP53, BRCA1/2,* and *STK11*. Other notable mutual exclusivities included *PIK3R1, FGF23, CCND2, MYC, KDM5A,* and *GATA3*.

*AKT1* was significantly co-altered with *NF1* and *CDH1*, an association which has been previously described in ILC^29^. There were several shared co-occurrence patterns with *PIK3CA*, including *MAP3K1*, which may reflect the increased prevalence of both *PIK3CA* and *AKT1* in ER+/HER2- tumors*. AKT1* was mutually exclusive with *ERBB2 and MYC*.

*PTEN* was significantly co-altered with *FAS* (also located on 10q23) as well as the tumor suppressors *TP53* and *RB1*. *PTEN* is known to be an essential mediator of Fas-mediated apoptosis, and *PTEN* loss can lead to impaired apoptosis and increased cell survival^30^. Combined *PTEN* and *TP53* loss is known to be associated with poor therapeutic response and poor prognosis, particularly in triple negative breast cancers^31,32^. Notable mutually exclusivities with *PTEN* included *ERBB2, GATA3,* and *ESR1*.

Co-occurrence analyses were also performed on VUS in the three genes of interest, which showed weakly significant results (Supplementary Figure S1; Supplementary Tables S7-S9).

### Functional characterization of clinical PTEN alterations

Among the more than 1,700 clinical *PTEN* variants identified in our cohort, a notable proportion are variants of uncertain significance with unclear functional implications. We leveraged large-scale experimental data sets previously generated in cell models to explore the functions of clinical *PTEN* variants in our cohort. Specifically, we examined two published deep mutational scanning (DMS) data sets characterizing the effects of single amino acid substitutions on *PTEN* abundance (protein expression) or function (effects on lipid phosphatase activity as measured by PIP_3_ levels)^18–20^. The original published data sets generated abundance scores for 4,547 missense or nonsense *PTEN* variants and fitness scores for 6,904 missense or nonsense *PTEN* variants.

Our clinical cohort included 610 single amino acid substitution *PTEN* variants (494 missense and 116 nonsense). Of these, 339 (55.6% of all *PTEN* variants) had an available abundance score and 536 (87.9%) had an available fitness score; 298 (48.9%) variants had both abundance and fitness scores available. Abundance and fitness scores for each missense and nonsense *PTEN* variant in our clinical cohort are collated (Supplementary Table S10).

As expected, nonsense variants exhibited statistically significantly lower abundance and fitness scores compared to missense variants (*p*=1.12x10^-28^ and *p*=4.93x10^-25^ for abundance and fitness scores, respectively; Figure 4A). Nearly all nonsense variants with available DMS data were appropriately classified as pathogenic by both abundance score (n = 49/49; 100%) and fitness score (n = 88/92; 95.7%). However, there was significant variation in the functional classifications assigned to individual missense variants. By abundance score, 148 (51.0% of missense variants with available abundance score) were pathogenic and 142 (49.0%) were non-pathogenic; by fitness score, 231 (52.0% of missense variants with available fitness score) were pathogenic and 213 (48.0%) were non-pathogenic. For missense variants with both abundance and fitness scores available (n = 261), we compared the functional classifications assigned by each assay (Figures 4B and 4C). Notably, 31.8% (83/261) of missense variants were classified as non-pathogenic by both assays, highlighting a substantial proportion of missense *PTEN* variants that do not contribute to pathogenicity. Meanwhile, 35.6% (93/298) were concordantly classified as pathogenic by both assays. The remaining one-third of missense variants had discordances in functional classification (85, 32.5%), exhibiting either loss of abundance without loss of phosphatase activity (39, 14.9%) or loss of phosphatase activity without loss of abundance (46, 17.6%). These discordances likely reflect the distinct phenotypic effects of mutations at different amino acid residues.

**Figure 4.**
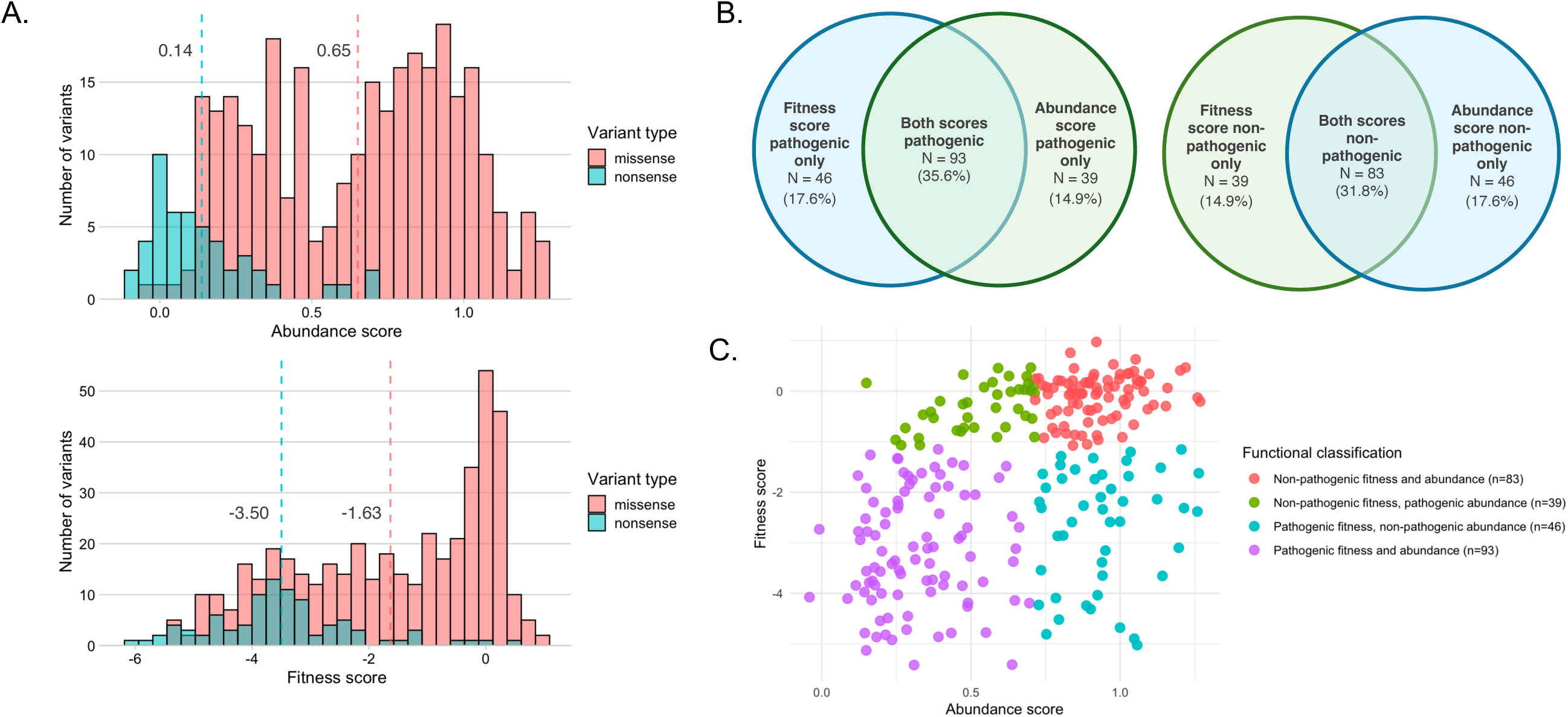
Deep mutational scanning data for missense and nonsense *PTEN* variants. **A)** Distribution of abundance and fitness scores among missense versus nonsense variants in the cohort, from Matreyek et al 2021 and Mighell et al 2018. Mean abundance and fitness scores for nonsense variants are significantly lower than for missense variants (mean scores for each group are represented by dashed lines; *p*=1.12x10^-28^ and *p*=4.93x10^-25^, respectively). **B)** Set diagrams depicting degree of concordance in functional classification by abundance versus fitness score, for missense variants only. **C)** Scatterplot of abundance and fitness scores for missense variants only, colored by concordance in functional classification between the two assays.

## Discussion

Here we report the largest landscape study of *PIK3CA*, *AKT1,* and *PTEN* genomic alterations in breast tumors to date, encompassing 29,157 alterations across the three genes. The frequency and distribution of *PIK3CA* and *AKT1* alterations in this cohort was consistent with prior reports^11–13,22,33^. Interestingly, the frequency of *PTEN* alterations was higher (13.5%) than in prior reports of *PTEN* alterations in breast cancer (≤10%) ^12,13^, which could be attributable to the detection of additional *PTEN* variants compared to prior studies which used targeted sequencing of certain exons.

The most common *PIK3CA* variants we observed were H1047R, E545K, E542K, N345K, and H1047L, consistent with previous reports^34,35^. While hotspot mutations in *PIK3CA* are well-established as predictors of response to AKT pathway inhibitors, the roles of non-hotspot mutations are variably understood. A subset of these rare variants were included in the companion diagnostic used in the SOLAR-1 trial^5,35^, while others including the fourth most common mutation in our data set (N345K)^36,37^ have been associated with alpelisib sensitivity in preclinical studies. We found that 16.2% of *PIK3CA*-altered samples had multiple *PIK3CA* alterations, similar to previous reporting^38^, which have been shown to result in increased PI3K activity and increased sensitivity to PI3Kα inhibitors. *AKT1* E17K accounted for over two-thirds of variants in the gene, leaving a notable minority of non-hotspot alterations whose biological and clinical implications remain to be clarified. One study found that a subset of non-E17K missense mutations in the long tail of *AKT1* and *AKT2* led to increased PI3K signaling and sensitized tumors to AKT inhibition; additionally, a group of in-frame duplications led to distinct structural changes, AKT activation, and hypersensitivity to ATP-competitive but not allosteric AKT inhibitors^39^. Further clarification of mutant-specific effects on pathway activity and mutant-specific therapeutic vulnerabilities represent an important outstanding area of investigation. We found that ER-/HER2- cases had a higher proportion of non-hotspot *PIK3CA* variants than other receptor subtypes. However, given negative results from several prior trials of PI3K pathway inhibition in ER-/HER2- breast cancer^40–43^, it remains to be clarified whether hotspot and/or non-hotspot *PIK3CA* variants predict for PI3K/AKT inhibitor response in triple negative disease.

Our demographic analyses revealed that *PIK3CA* alterations were less common in patients with African ancestry than other groups, among all receptor subtypes and in ER+/HER2- disease alone, which has been demonstrated in smaller prior studies^44–47^. Here we additionally found no difference in the prevalence of *AKT1* and *PTEN* variants across ancestries, suggesting that ancestry-based differences in *PIK3CA* prevalence are gene-specific rather than affecting the PI3K pathway globally. This could be due to differential tumor biology and mechanisms that may limit in African American patients the acquisition of *PIK3CA* mutations which are early clonal events, and highlight one biological explanation for lower representation of African American patients in *PIK3CA* biomarker-based targeted therapy trials. We also found that *PIK3CA* prevalence was significantly higher in patients with South Asian ancestry and ER+/HER2-disease, underscoring the need for representative patient enrollment in biomarker-based clinical trials.

Our gene association analyses corroborated prior findings that *PIK3CA*, *AKT1*, and *PTEN* pathogenic alterations are strongly mutually exclusive with each other^11,28,35^, likely reflecting redundancies in PI3K pathway activation between these genes. Several significant co-occurrences involving *PIK3CA* and *AKT1* were consistent with the literature and with their prevalence in certain histopathologic subtypes. These included previously recognized associations with *MAPK1* and *CDH1/NF1*, which are known to be enriched in ER+ disease and ILC, respectively. *MAPK1* loss-of-function variants occurring in *PIK3CA*-mutated cell lines have also been shown to result in resistance to PI3Kα inhibitors and AKT inhibitors^27^, suggesting their potential value as a predictive biomarker for AKT pathway inhibitor resistance. We identified a novel co-occurrence tendency between *PIK3CA* and *SOX2*, a combination which has been implicated in the pathogenesis of head and neck cancers but is not well-described in breast cancers. *PIK3CA* and *AKT1* tended to be mutually exclusive with tumor suppressors such as *MYC*, while *PTEN*-altered samples were enriched for mutations in other tumor suppressors such as *FAS* and *TP53*, a combination which is recognized as a poor prognostic factor especially in triple negative disease.

The functional characterization of *PTEN* VUS remains a challenge in clinical genomics, as accurate classification can directly impact patient management. Our analysis highlights the value of large-scale experimental datasets, such as DMS, in resolving the pathogenicity of these variants. Notably, we identified a substantial proportion of missense *PTEN* variants (31.8%) that were consistently classified as non-pathogenic by both abundance and fitness-based assays, underscoring the importance of functional data in refining variant interpretation and potentially reducing the burden of uncertainty in clinical decision-making. However, discordances in nearly one-third of missense variants suggest that a substantial proportion of mutations may not predict straightforward loss of function and therefore warrant further mechanistic investigation in the laboratory as to if they predict for AKT inhibitor sensitivity. Expanding and refining such datasets will be critical for improving precision in molecular diagnostics and therapeutic guidance.

Our study has limitations including the lack of available ER status on a substantial subset of patients, which may limit the interpretation of the related analyses, though the mutational distributions that we observed across receptor subtypes were consistent with prior reports. Samples are restricted to patients residing in the United States, which could limit the generalizability of the ancestry-related findings.

In conclusion, we have leveraged a large CGP dataset of >51,000 breast tumors to generate insights on the clinicogenomic landscape of an important biomarker triad in breast cancer. We identify a wide spectrum of alterations in *PIK3CA/AKT1/PTEN* encompassing well-established pathogenic variants as well as rare variants with unknown clinical significance. The prevalence of non-hotspot mutations, some of which are known to mediate sensitivity to older PI3K inhibitors, supports the use of CGP over smaller gene panels for identifying potentially actionable variants. Importantly, further preclinical and clinical studies are needed to clarify the roles of lesser-known variants as predictive biomarkers for the expanding collection of PI3K pathway inhibitors. Ancestry-based differences in *PIK3CA* prevalence are significant and should be taken into consideration in clinical studies of PI3K inhibition. As shown here, DMS data sets can be leveraged to generate insights into the functional implications of rare clinical variants.

## Data Availability

All relevant data are provided within the article and its accompanying Supplementary Data.

---

**Supplementary Figure S1.**
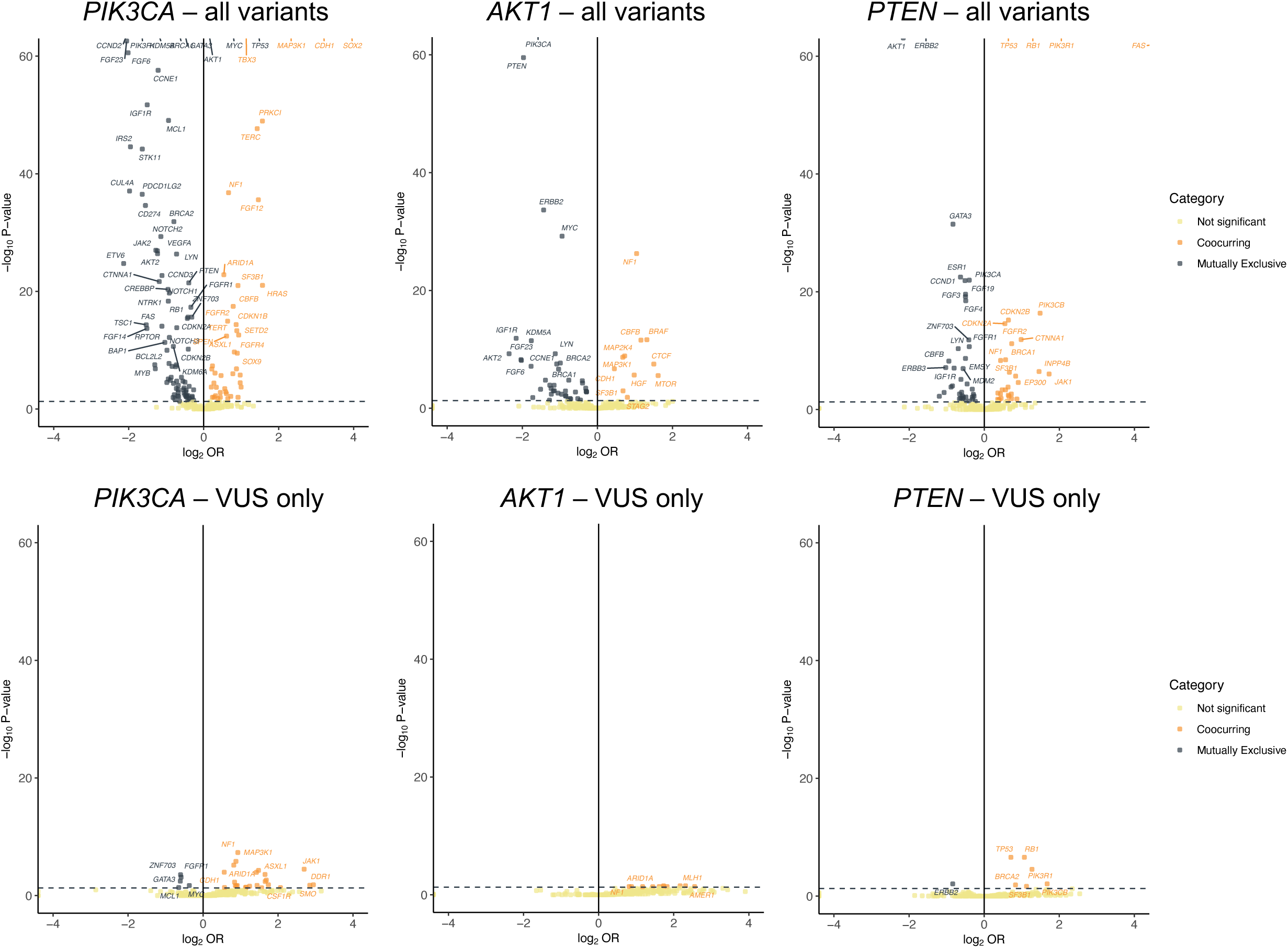
Co-occurrence and mutual exclusivity analyses of all variants (pathogenic + VUS; top) and VUS only (bottom) in *PIK3CA*, *AKT1*, and *PTEN*. Y-axis is capped at p-value of 10E-60.

